# Evaluation of the impact of concentration and extraction methods on the targeted sequencing of human viruses from wastewater

**DOI:** 10.1101/2024.01.18.24301434

**Authors:** Minxi Jiang, Audrey L.W. Wang, Nicholas A. Be, Nisha Mulakken, Kara L. Nelson, Rose S. Kantor

**Affiliations:** Department of Civil and Environmental Engineering, University of California, Berkeley, CA, USA; Physical and Life Sciences Directorate, Lawrence Livermore National Laboratory, Livermore, CA, USA; Computing and Global Security Directorates, Lawrence Livermore National Laboratory, Livermore, CA, USA

**Keywords:** Targeted sequencing, probe-capture enrichment, human virus, wastewater-based surveillance, wastewater-based epidemiology, virus concentration, nucleic acid extraction

## Abstract

Sequencing human viruses in wastewater is challenging due to their low abundance compared to the total microbial background. This study compared the impact of four virus concentration/extraction methods (Innovaprep, Nanotrap, Promega, Solids extraction) on probe-capture enrichment for human viruses followed by sequencing. Different concentration/extraction methods yielded distinct virus profiles. Innovaprep ultrafiltration (following solids removal) had the highest sequencing sensitivity and richness, resulting in the successful assembly of most near-complete human virus genomes. However, it was less sensitive in detecting SARS-CoV-2 by dPCR compared to Promega and Nanotrap. Across all preparation methods, astroviruses and polyomaviruses were the most highly abundant human viruses, and SARS-CoV-2 was rare. These findings suggest that sequencing success can be increased by using methods that reduce non-target nucleic acids in the extract, though the absolute concentration of total extracted nucleic acid, as indicated by Qubit, and targeted viruses, as indicated by dPCR, may not be directly related to targeted sequencing performance. Further, using broadly targeted sequencing panels may capture viral diversity but risks losing signals for specific low-abundance viruses. Overall, this study highlights the importance of aligning wet lab and bioinformatic methods with specific goals when employing probe-capture enrichment for human virus sequencing from wastewater.

**Synopsis:** Four concentration/extraction methods combined with probe-capture sequencing of human viruses in raw wastewater were compared. Innovaprep ultrafiltration with solids removal had the best performance for human virus detection sensitivity, richness, and recovery of near-complete genomes.

## 1. Introduction

Wastewater-based epidemiology (WBE), previously employed for monitoring enteric viruses like polio ^1^, has been widely applied during the COVID-19 pandemic. In 2020, the US Centers for Disease Control and Prevention (CDC) launched the National Wastewater Surveillance System (NWSS), to build and coordinate the capacity for WBE as a component of the nationwide monitoring of SARS-CoV-2 ^2^. Subsequently, groups around the world have expanded WBE to include PCR-based monitoring of known seasonal respiratory viruses including RSV and Influenza A and B, and new PCR panels are expected to contribute to CDC NWSS ^3^.

Unlike PCR-based virus quantification, sequencing of viruses in wastewater has the potential to monitor many human viruses at the genome level simultaneously. Reference-based amplicon sequencing using tiled panels such as ARTIC SARS-CoV-2 ^4^, ARTIC HAdV-F41 ^5^, Swift Normalase™ Amplicon Panel ^6^, or targeted amplicons like those for the VP1 or VP4 regions of enterovirus ^7, 8^, have enabled subtyping and tracking of circulating variants and strains, providing evidence that wastewater data aligns with available clinical data ^4, 5^. However, amplicon-based sequencing is limited in its ability to detect novel viruses, due to the challenges of degenerate primer design and multiplexing. In contrast, deep untargeted sequencing offers a comprehensive view of viral diversity in wastewater ^9–11^, but human viruses constitute a minimal fraction of the microbial nucleic acids present in wastewater, approximately 0.011% of unique reads ^10^ or 0.1% of the assembled contigs ^11^. To increase sequencing coverage of human viruses and to allow the detection of divergent or novel viruses in wastewater, probe-capture enrichment panels have been adopted from clinical research ^12^. Here, probes hybridize to DNA targets in a sample, allowing downstream separation of targets from background DNA. Because probe hybridization allows more mismatches than primer binding during PCR, more divergent sequences may be enriched by probe capture, potentially including novel relatives of known viruses. Recent studies that have applied virus probe-capture panels to wastewater-derived samples reported an increase in the proportion of viral reads up to 81% compared to untargeted sequencing ^13^. Although probe-capture-based sequencing enriched human viruses, most of the recovered viral content (> 80%) still consisted of bacteriophages and plant viruses ^14, 15^. These findings indicate that probe capture panels are still limited in their ability to enrich target sequences in samples with large amounts of background/non-target sequences, suggesting that the choice of upstream sample processing method may affect the detection of human viruses.

Prior to the COVID-19 pandemic, sequencing-based wastewater virus studies relied on large-volume time-intensive methods that had initially been developed to culture infectious viruses (e.g., polyethylene glycol precipitation, skim milk flocculation, ultracentrifugation, and membrane filtration). Multiple studies reported that the choice of concentration method influenced the resulting virus profiles by untargeted sequencing ^9, 16^ and few studies reported any sequences from enveloped viruses. During the pandemic, the demand for rapid routine monitoring of SARS-CoV-2 led to the development and wider adoption of streamlined concentration/extraction methods with lower sampling volumes, ending with qPCR or digital PCR quantification^17, 18^. These methods included size separation (e.g., Innovaprep ultrafiltration pipette, centrifugal ultrafiltration), capture based on virus surface characteristics (e.g., Nanotrap beads, electron-negative HA membrane), and direct nucleic acid extraction (e.g., Promega Wizard Enviro large-volume extraction, or extraction of wastewater solids after centrifugation). These routine monitoring methods were also used to obtain SARS-CoV-2 RNA for sequencing, with varying success ^19–22^ and later extended for detection of a wider spectrum of viruses ^14, 15, 23–25^. To date, few studies have directly compared the effects of different methods on the success of virus probe-capture enrichment sequencing. McCall et al. (2023) compared methods with very different sampling volumes (300 μL for direct extraction and 50 mL for HA filtration) and suggested that direct extraction may yield a lower equivalent volume of viruses in the final extracted nucleic acid compared to pre-filtered samples^23^. Spurbeck et al (2023) indirectly compared five wastewater virus concentration/extraction methods, but each was applied to wastewater samples from a different location(s). They found that Innovaprep ultrafiltration yielded the highest virus sequence recovery in untargeted RNA sequencing, although most sequences corresponded to bacteriophage ^24^. These findings highlight the potential impact of concentration/extraction methods on targeted sequencing of diverse viruses, but direct comparisons and analysis of potential biases from concentration methods on sequencing performance are needed, especially for targeted sequencing using probe-capture panels.

In this study, four wastewater virus concentration/extraction methods were selected based on their ongoing use in wastewater surveillance efforts, and the success of probe-capture enrichment sequencing was compared for each method. The wastewater input volume was held constant, and the resulting nucleic acids were enriched using the Illumina virus surveillance panel (VSP). The evaluation of methods performance included total nucleic acid quality, unique sequence output, taxonomic composition, richness, recovered genome completeness, and sensitivity comparisons between sequencing and dPCR. Ultimately, these findings improve our understanding of wet lab approaches and their compatibility with virus probe-capture enrichment and sequencing, informing tailored responses to emerging viral threats.

## 2. Materials and Methods

### 2.1 Sample collection

Influent wastewater was collected as 24-hour composite samples on three dates: March 1st, April 19th, and April 26th, 2023, from the EBMUD wastewater treatment plant (Alameda County, CA). This facility serves approximately 700,000 people, receiving domestic and industrial wastewater. On each date, the sample was transported to the laboratory on ice, and twelve 40 mL aliquots were prepared. Bovine coronavirus (BCoV) was added to each tube as a sample processing control to assess viral RNA recovery. First, one vial of BCoV (Merck) vaccine powder was resuspended in 2 mL 0.1 mM Tris-Ethylenediaminetetraacetic acid (TE) buffer and diluted 10-fold. Each wastewater aliquot was spiked with 50 µL BCoV solution and incubated overnight at 4℃.

### 2.2 Concentration and extraction

Four concentration and extraction methods (described below) were employed in this study: Innovaprep pipette concentration (IP method), Nanotrap bead concentration (NT method), Promega large-volume direct extraction (PMG method), and pelleted solids direct extraction (Solids method). Each method was performed on three 40-mL aliquots of wastewater per sample date, alongside a negative control consisting of 40 mL 1x phosphate-buffered saline (PBS) solution (Table S1). All methods resulted in 100 µl purified total nucleic acid (TNA).

In the first two methods, separate concentration and extraction steps were applied. In the IP method, 400 µL of 5% Tween 20 was added to the wastewater sample and mixed by inversion, followed by centrifugation at 7000 x g for 10 min. The supernatant was ultrafiltered using the automatic HF Concentration Pipette (Innovaprep CP-Select™) and eluted with the elution fluid (Innovaprep) to produce the viral concentrate (ranging from 160 to 882 μL, Table S1). TNA was then extracted from up to 200 µL of viral concentrate using the Allprep PowerViral DNA/RNA kit (Qiagen) and eluted in 100 μL, following the manufacturer’s liquid sample extraction protocol. The NT method followed the Nanotrap® Microbiome A Protocol, compatible with AllPrep PowerViral DNA/RNA Kit (APP-091 December 2022). Briefly, 115 µL of Nanotrap® Enhancement Reagent 2 (ER2) and 600 µL of Nanotrap Microbiome A Particles (Ceres Nanosciences) were sequentially added to each sample, followed by mixing and incubation. The beads were separated from the solution on a magnetic rack and resuspended in 1 mL of molecular-grade water, followed by another separation using the magnetic rack. The beads were then mixed with 600 µL of preheated PM1 + Beta-mercaptoethanol solution from the Allprep PowerViral kit, and the mixture was heated at 95℃ for 10 min to release nucleic acids. Beads were removed using the magnetic rack and the supernatant was used for subsequent extraction steps using the Allprep PowerViral DNA/RNA kit liquid protocol, resulting in 100 µL of final TNA.

The other two methods were direct extractions from either total wastewater or pelleted solids. The PMG method used the commercial kit from Promega (Wizard® Enviro Total Nucleic Acid) following the manufacturer’s protocol. Briefly, 0.5 mL of protease was added to each 40 mL wastewater sample and incubated for 30 minutes. After centrifugation at 3000 x g for 10 min, binding buffers and isopropanol were added to the resulting supernatant before passing it through the PureYield™ binding column. The bound nucleic acids were washed and then eluted in 1 mL of nuclease-free water. The eluted samples were further purified, concentrated, and eluted using the PureYield™ Minicolumn, resulting in a final total nucleic acid volume of 100 µL. In the Solids method, the 40 mL wastewater sample was centrifuged at 20,000 x g for 10 minutes to pellet the solids. Total nucleic acid was then extracted from 0.2 g (wet weight) of solid pellets using the Allprep PowerViral DNA/RNA extraction kit. This followed the manufacturer’s solids extraction protocol, which included a 10-minute bead-beating step after the addition of PM1 and Beta-mercaptoethanol solution. The final extracted TNA were eluted in 100 µL of nuclease-free water.

DNA and RNA concentrations were quantified using the Qubit 1X dsDNA HS Assay (Fisher Scientific) and Qubit RNA HS Assay (Fisher Scientific), respectively. Aliquots of all extracts were stored at -20℃ and quantified by dPCR within one week and at -80℃ for subsequent sequencing library preparation.

### 2.3 Digital PCR quantification of SARS-CoV-2 in the extracted total nucleic acid

Digital PCR was performed on the QIAcuity Four Platform Digital PCR System (Qiagen). The details of SARS-CoV-2 and BCoV assays’ primers, probes, and thermal cycling conditions are summarized in Table S2a. The reaction mixtures were prepared using the QIAcuity OneStep Advanced Probe Kit (Qiagen) and loaded onto either 8.5k 24-well or 26k 24-well nanoplates (Qiagen). Details of reaction mixture composition and volumes were summarized in Table S2b. The positive control was linearized gene plasmids from Integrated DNA Technologies, and the negative control was nuclease-free water. See Figure S1 for examples of the partition fluorescence plots of positive and negative controls. The number of valid partitions ranged from 7,920 to 8,269 per well for 8.5k plates and 12,548 to 25,493 per well for 26k plates. Data were analyzed using the QIAcuity Suite Software V1.1.3 (Qiagen, Germany) with automated settings for threshold and baseline, followed by manual inspection. Results were plotted using a customized Python script. dMIQE checklists ^26^ are provided in Table S3. The operational limit of detection was treated as ≥3 positive partitions per well.

### 2.4 Library preparation and targeted sequencing

Before library preparation, DNA and RNA quality were measured by Fragment Analyzer with the default HS NGS Fragment 1-50 kb assay and Bioanalyzer (Agilent 2100) with the Agilent RNA 6000 Pico RNA assay, respectively. Library preparation followed the Illumina RNA Prep with Enrichment kits with modifications to total input (Illumina, San Diego, CA, USA). In brief, the mixture of purified DNA and RNA from samples collected on April 19 and April 26 was diluted with nuclease-free water such that the final concentration of RNA was ≤ 100 ng/µl. Dilution was not conducted for IP and NT samples due to the low RNA concentrations. The DNA and RNA from samples collected on March 1 were used directly as the input for library preparation without dilution for all concentration/extraction methods (Table S1). Next, 8.5-µL of each sample was denatured followed by first- and second-strand DNA synthesis. Tagmentation of the total enriched double-stranded cDNA was performed using bead-linked transposons (BLT), and adapter sequences were added at the same time. The resulting fragments were purified and amplified to add index sequences. Libraries were quantified using the Qubit dsDNA broad-range Assay Kit. Enrichment was performed with the Illumina VSP Panel by pooling 200 ng of each library from three biological replicates into hybridization reactions. This step was followed by bead-based capture of hybridized probes, amplification, clean-up, and quantification of the final enriched library. After library preparation, all enriched samples were pooled in equimolar ratios and sequenced on one lane of Illumina Novaseq 6000 SP 150PE.

### 2.5 Bioinformatics Analysis Pipeline

Sequence data were quality trimmed using BBduk ^27^ to remove adaptors and filter out low-quality reads and short reads. Seqkit was used to deduplicate reads and summarize unique reads ^28^ (Figure 1b). Before taxonomy classification, human reads were filtered using bowtie2 (v2.5.1) ^29^ by mapping to GRCh38.p14 (RefSeq GCF_000001405.40) and CHM13v2.0 (RefSeq GCF_009914755.1). The remaining non-human unique reads were classified by Centrifuge (1.0.4) ^30^ and Recentrifuge ^31^ using a decontaminated version of NCBI-nt database (NCBI release date June 5, 2023). A minimum hit length (MHL) threshold of 15 was employed for Centrifuge. An MHL threshold of 40 was subsequently applied in Recentrifuge for downstream analysis. After classification, all viral reads were extracted from each sample using rextract and viral sequence similarities between samples were compared using MASH ^32^. Pairwise Mash distances were calculated for the construction of the PCoA plot using the sklearn.decomposition PCA package in Python. A PERMANOVA test with 999 permutations was performed using the vegan package (2.6.4) in R ^33^. One sample (PMG_426_2) displayed distinct sequence properties from the other two biological replicates in the original PCoA (Figure S2) and yielded unexpectedly low unique read counts (Table S1), likely due to unsuccessful enrichment during the library preparation. This sample was excluded from all sequencing analyses. To precisely identify SARS-CoV-2 reads during the sampling period, unique reads classified by Centrifuge at the species level as severe acute respiratory syndrome-related coronavirus (taxID: 694009) were extracted and mapped to references from the GISAID database ^34^ downloaded on January 2, 2024 (Table S4). The references comprised 463 complete genome sequences with high coverage and collection dates ranging from January 1, 2023, to May 31, 2023. The mapped reads were subjected to additional filtering using reformat.sh from BBduk ^27^. Mapped reads with fewer than 5 mismatches were considered as classified SARS-CoV-2 reads.

**Figure 1.**
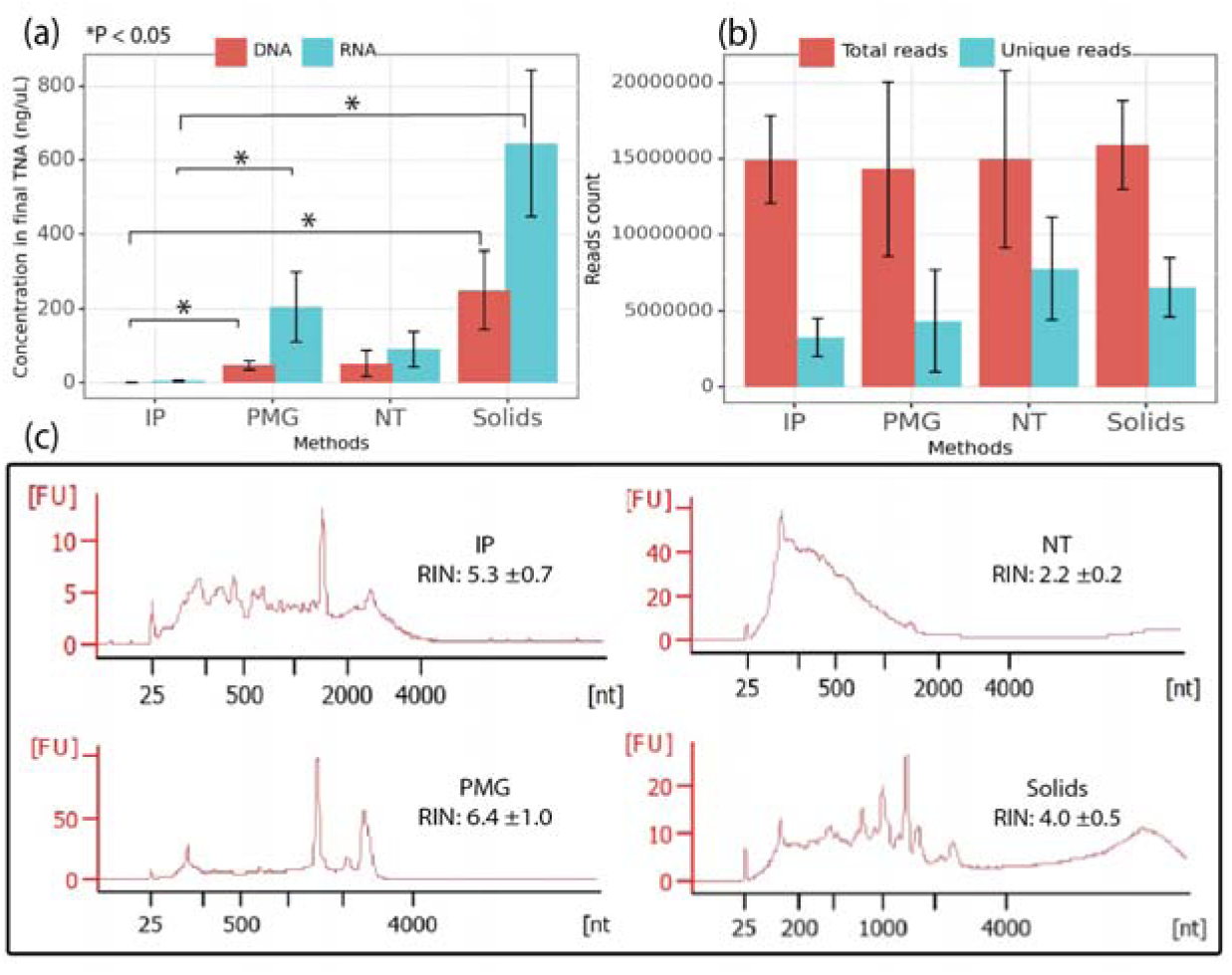
Nucleic acids and unique read counts by sample processing method. (a) Averaged concentrations of extracted DNA and RNA produced by each method (n=9 samples per method); (b) Averaged raw read counts and counts of unique reads after QC trimming and deduplication in each method (n = 9 samples for IP, NT, and Solids, n = 8 for PMG); (c) Representative RNA fragment size distribution and average RNA integrity number (RIN) for each method. Note that samples were diluted before fragment analysis (IP: undiluted, NT: 25x, PMG: 25x, Solids: 200x), so y-axes are not comparable.

To determine putative virus host assignments for each read (Figure 2c and 2d), the NCBI taxonomy database ^35^, which includes virus host information, was queried with the NCBI taxID of the best hit given by Recentrifuge, and the results were manually inspected (see SI methods for details). The comparison of DNA and RNA viruses was conducted at the virus kingdom level based on taxonomic classification results. To focus specifically on human viruses, the identification of all human viruses (section 3.3) occurred at the species level based on NCBI taxonomy ^35^. Species-level richness was determined by counting the unique human virus species, with a cutoff of > 10 classified reads applied to discard low-abundance viruses (Figure S3a and 3b). Some reads were assigned to species-level NCBI taxIDs that paradoxically lacked clear species-level taxonomy names in the database. To make this apparent, these species are displayed with “unclassified” appended to the species name (see SI methods for details).

**Figure 2.**
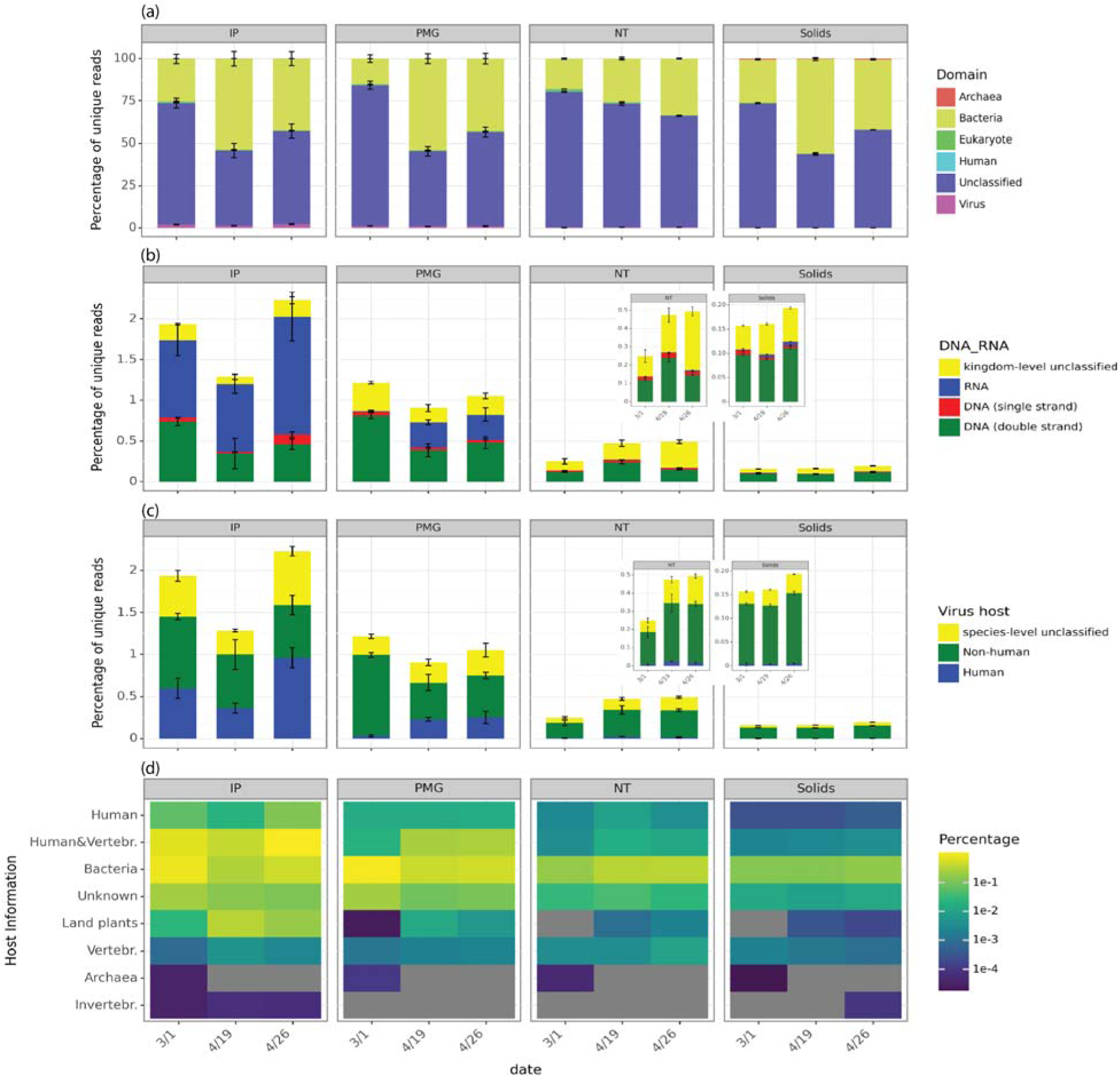
Taxonomic profiles of reads and virus hosts differed by method. (a) Domain-level classification of unique reads by Recentrifuge, with samples collected on three sampling dates and processed by four methods (n=3, except Promega 4/26). “unclassified” is the sum of reads discarded by Recentrifuge without taxonomic classification and thos classified as “Root” but without a domain-level classification. “Human” represented unique reads mapped to two downloaded human genomes (see methods); (b) Percentages of unique reads identified as RNA, double-strand DNA, and single-strand DNA viruses based on kingdom-level virus classification; (c) Percentages of unique reads identified as virus species linked to human and non-human hosts in NCBI or for which species-level taxonomy was not determined; (d) Percentages of unique viral reads associated with different host categories in the NCBI Virus database. Note that “human” in (c) encompasses the categories “human & vertebrates” and “human” in (d). In (d), reads assigned to BCoV were subtracted from counts of reads assigned t “human & vertebrates” and are not displayed.

To maximize the recovery of near-complete viral genomes for wet lab method comparison, all unique reads were separately assembled using SPAdes with the -meta option (v3.15.5) ^36^. All virus scaffolds identified by VirSorter2 ^37^ were further subjected to quality filtering, requiring a length of > 1000 bp and an average coverage of > 10x. These filtered assemblies were then subjected to BLASTn search against the NCBI nt virus database, with stringent quality filters applied: > 80% identity, > 90% alignment/query length, and an e-value < 1E-8. The best hit based on bitscore was retained for each assembled scaffold and information including virus name, taxID, genome completeness, and genome length was retrieved from NCBI via the dataset and dataformat functions ^38^. Hit genomes were retained only if complete, and assembled genomes were used for further analysis if the alignment length was > 70% of the complete hit genome, indicating the assembly of a near-complete genome from wastewater (Figure S4b).

Scaffolds representing near-complete genomes for JC polyomavirus were collected for phylogenetic analyses. Potential assembly errors were inspected by mapping reads to assembled scaffolds using bowtie2 and visualizing with the Integrative Genomics Viewer (IGV) ^39^. No assembly errors were detected, and representative mappings are shown in Figure S5. Given the circularity of the JC polyomavirus genome, assemblies were also examined in Geneious ^40^, and repeated regions at the beginning and the end of the sequences were trimmed before the alignment (Figure S6). Multiple sequence alignment was performed by MUSCLE ^41^ with all trimmed scaffolds, all JC polyomavirus reference genomes from NCBI GenBank released within two years (n=39), and the best-hit results from BLASTn for each scaffold. The alignment was inspected in Geneious to identify a common starting point, and all 10 scaffolds were recircularized to this point. The recircularized scaffolds were queried against NCBI again to identify new best-hit reference genomes, which may have changed due to genome curation. The final dataset included these curated genomes, new best-hits, and the 39 JC polyomavirus references. Alignment was performed with MUSCLE followed by GBlocks ^42^ to identify informative regions, and MEGA 11.0 ^43^ was used to generate and visualize the final maximum likelihood tree using the Tamura Nei model with 100 bootstrap replicates.

### 2.6 Statistical analysis and data availability

The normality of data was assessed using the Shapiro-Wilk test. Statistical differences between concentration and extraction methods were evaluated using the Kruskal-Wallis test, followed by post-hoc pairwise Dunn’s test. All statistical tests were performed using the Python package scipy.stats, and significance was determined at a 95% confidence interval (p < 0.05). Sequencing data for this project for this project has been deposited in the NCBI Sequence Read Archive (SRA) under accession number: SUB13892842 and Bioproject ID: PRJNA1047067. The processed data, reproducible code, and the analysis workflow are available at https://github.com/mj2770/Wastewater-virus-surveillance-.

## 3. Results and Discussion

In this study, wastewater influent was collected from a single WWTP on three dates, and viruses were concentrated and extracted by four methods: IP method (Innovaprep ultrafiltration of liquid portion paired with a small-volume extraction kit), NT method (Nanotrap beads-based affinity capture performed on total influent paired with small-volume extraction kit), PMG method (Promega large volume direct extraction), and Solids method (centrifugation paired with small-volume extraction kit). The resulting 36 samples (12 samples in biological triplicate) were processed using the virus surveillance panel (VSP) from Illumina using probe-capture enrichment. Following the initial analysis, an outlier sample was identified, indicating unsuccessful library preparation (see Methods), and this sample was excluded from all analyses.

### 3.1 Sample quality and sequence data

The DNA and RNA generated using the four methods differed in concentration (Kruskal-Wallis test p = 2E-6 and 7E-7, respectively), fragment size distribution, and RNA integrity (ANOVA test p = 1E-13). The Solids method consistently resulted in yields that were higher than other methods for both DNA and RNA (Figure 1a), while the IP method, which includes a solids removal step, resulted in significantly lower total DNA and RNA yield compared to Solids and PMG (Figure 1a, IP v.s. Solids DNA p = 3E-7, IP v.s. PMG DNA p = 0.02, IP v.s. Solids RNA p = 3E-7, IP v.s. PMG RNA p = 0.004). All methods yielded a higher concentration of RNA than DNA, but the resulting ratios of RNA:DNA varied significantly (Kruskal-Wallis test p = 0.002) across methods from 2.0 ± 0.7 (for NT) to 4.3 ± 1.6 (for PMG). Unlike the other methods, shorter RNA fragments were observed with the NT method and 16S rRNA and 23S rRNA were absent, perhaps accounting for the low RNA:DNA ratio. The lack of ribosomal RNA may be due to the exclusion of bacteria by the nanotrap hydrogel particle shells, which have specific pore sizes and are chemically modified to prevent the entry and capture of large or non-targeted particles ^44^. Although viral RNA integrity is not discernible from the RNA Integrity Number (RIN) alone, the highest RIN was observed with the PMG method (6.4 ± 1.0, Figure 1c), which suggested more intact prokaryotic RNA was preserved with the PMG method.

After sequencing 36 samples, a total of 535 million reads were generated, averaging 14.86 ± 4.46 million reads per sample (Figure 1b), and the removal of PCR duplicates reduced read counts by over 50% for all samples. As the IP method produced the lowest RNA and DNA input concentrations, it was not surprising that after deduplication these samples also retained significantly fewer unique reads (3.3 ± 1.3 million, Figure 1b) compared to samples from the Solids and NT methods (IP vs. NT p = 0.005, IP vs. Solids p = 0.04). Nonetheless, the count of unique reads was not clearly related to the DNA and RNA concentrations, perhaps due to the dilution of nucleic acids (Table S1) before library preparation, and the multiple amplification and equimolar pooling steps during library preparation.

### 3.2 Taxonomic classification and virus composition similarity

Over 40% of unique reads were not taxonomically classified by Recentrifuge at the Domain level with the selected Minimum Hit Length (MHL) across all methods, and most classified reads were assigned to the domain Bacteria (ranging from 25.84 ± 6.81% to 40.88 ± 13.13%, Figure 2a). It is likely that a larger proportion of unique reads would have received an assigned taxonomy at a lower classification stringency; however, such low-confidence assignments have the potential to introduce substantial noise to downstream assessments. Future functionalization of these platforms will require tuning of these stringency thresholds for the desired application, balancing classification sensitivity with assignment confidence. These findings could also reflect the current limitations of reference-based classifiers^46^ and limited enrichment of targets using probe-capture, irrespective of the concentration and extraction methods employed.

The percentage of reads classified as viral ranged from 0.17 ± 0.02% (Solids) to 1.82 ± 0.46% (IP) of unique reads across different methods (Figure 2b), surpassing the reported < 0.011% in untargeted sequencing ^9^. The IP samples yielded significantly higher percentages of viral reads than Solids and NT (1.82 ± 0.46%, Figure 2b, IP vs. Solids p = 8E-7, and IP vs. NT p = 0.004), followed by the PMG samples (1.06 ± 0.18%, Figure 2b, PMG vs. Solids p = 0.002). Additionally, the IP method concentrated significantly more RNA viruses (Figure 2b) and viruses associated with human and/or vertebrate hosts than NT and Solids methods (0.64 ± 0.27% human viruses in total unique reads from IP, Figures 2c and 2d, IP vs. NT p = 0.002, IP vs. Solids p =1E-6). The IP and PMG methods incorporated a solids removal step after attempting to release solid-associated viruses by adding 5% Tween 20 ^45^ or protease, respectively ^46^. These steps not only prevent clogging during sample processing but also strike a balance between eliminating solid-associated non-viral microorganisms like bacteria and attempting to retain viruses. As a result, a notably lower ratio of classified bacterial reads to classified viral reads was observed in IP and PMG samples (25 ± 14:1 and 38 ± 24:1, respectively) in comparison to Solids and NT samples (241 ± 83 and 66 ± 12, respectively) (IP vs. NT p = 0.04; IP vs. Solids p =1E-5; PMG vs. Solids p = 0.0006). In NT and Solids samples, most viral reads were associated with bacterial hosts, based on the NCBI taxonomy database (Figure 2d). This finding is consistent with the high fraction of DNA viruses in those samples (Figure 2b), as most bacteriophages are DNA viruses ^47^.

To compare virus composition across the four sample preparation methods, reads classified as viral by Recentrifuge were extracted from each sample, and MASH ^32^ was used to assess pairwise sequence similarity. In a principal component analysis using MASH distances, triplicate samples clustered together (PERMANOVA test p = 0.985), while all samples were separated by concentration/extraction methods along PC1 (37.2% of the variation, Figure 3, PERMANOVA test p = 0.001). Specifically, IP- and PMG samples clustered together, while NT and Solids samples were distinct (Figure 3). The predominance of bacteriophage in both NT and Solids samples likely contributed to their differentiation from the other two methods. Samples were separated by sampling dates along PC2 (24.5% of the variation, Figure 3, PERMANOVA test p = 0.001), with samples from March 1, 2023, differing from those collected on April 19 and April 26. This differentiation was observed consistently across all four methods. These temporal shifts in virus composition may suggest a temporally variable metavirome composition in wastewater, potentially influenced by changes in circulating viruses ^8, 48, 49^ and changing wastewater conditions, such as flow rate, total suspended solids (TSS), total organic compounds (TOC), and the abundance of antagonistic microorganisms ^50, 51^.

**Figure 3.**
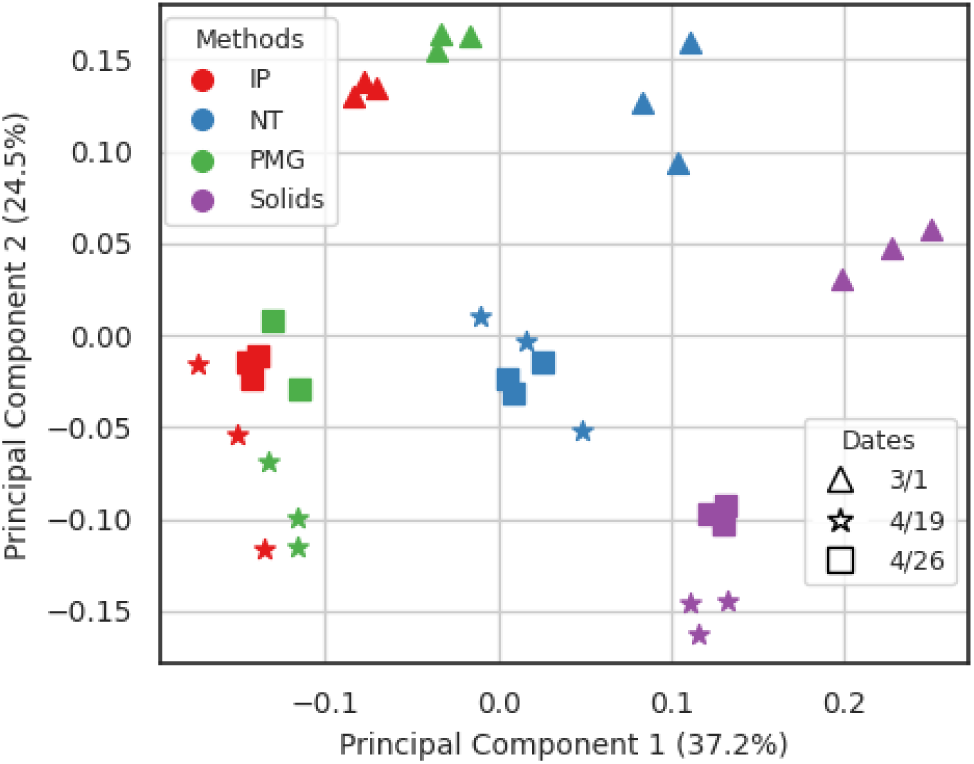
Viral sequence composition was influenced by wastewater virus concentration/extraction method and sample date. Principal component analysis (PCoA) plot was generated using the MASH distance, which was calculated based on sequence similarity among all reads classified as viral by Centrifuge. Different methods are represented by colors, and different sampling dates are represented by shapes.

### 3.3 Human virus species richness and composition

PMG and IP methods yielded higher species-level richness of total viruses detected with >10 reads (241 and 176 viruses, respectively) and human viruses (20 and 26 respectively) compared to NT and Solids (Figure S3a), although total read depth was similar for all samples (Figure 1b, p = 0.44). Thus, removing solids after releasing solid-associated viruses did not compromise the richness of detected human viruses. Conversely, including solids produced lower species-level diversity. Of the 66 virus “groups” of high public health significance listed as targets in the Illumina VSP panel (Table S5), IP samples detected members of 11 (Figure S3a). These included human coronavirus-OC43 (hCoV-OC43), adenovirus, astrovirus, aichivirus, enterovirus, norovirus, coxsackievirus, rotavirus, salivirus, and sapovirus, as well as mpox (Figure S3b), though the exact list of species and strains used by Illumina for probe design is proprietary; we note that enteroviruses are a diverse group which contains coxsackieviruses, while hCoV-OC43 is a sub-species level category.

All human virus species detected (>10 reads per species) in at least one sample were compared across the four methods (Figure 4). Some viruses were consistently detected by all methods, including human polyomavirus, mastadenovirus, mamastrovirus 1, and norwalk virus, which are known to be shed at high concentrations in human waste ^5, 9, 10, 13, 23, 48, 52–55^. RNA virus species, including severe acute respiratory syndrome-related coronavirus, sapporo virus, and enteroviruses were not detected in NT and Solids samples. Different trends were also observed among virus species within the same genus. For instance, human mastadenovirus B, D, and F were detected in all samples, while human mastadenovirus A, C, and E were not detected in certain samples (Figure 4). This variability suggested that related virus species may be differentially detected by different concentration methods. No arthropod-transmitted viruses (e.g., Dengue, Chikungunya), bloodborne viruses (e.g., Hepatitis virus and HIV), or hemorrhagic fever-related viruses (e.g., lassa mamarenavirus, junin virus, etc.) were detected, despite their inclusion in the probe panel. Mpox, detected intermittently in wastewater since the outbreak in 2022 ^56, 57^, was detected at low levels in IP, PMG, and NT samples. Overall, it seems reasonable to conclude that these results generally reflect a subset of current infectious diseases present in and absent from the San Francisco Bay Area at the time of sample collection.

**Figure 4.**
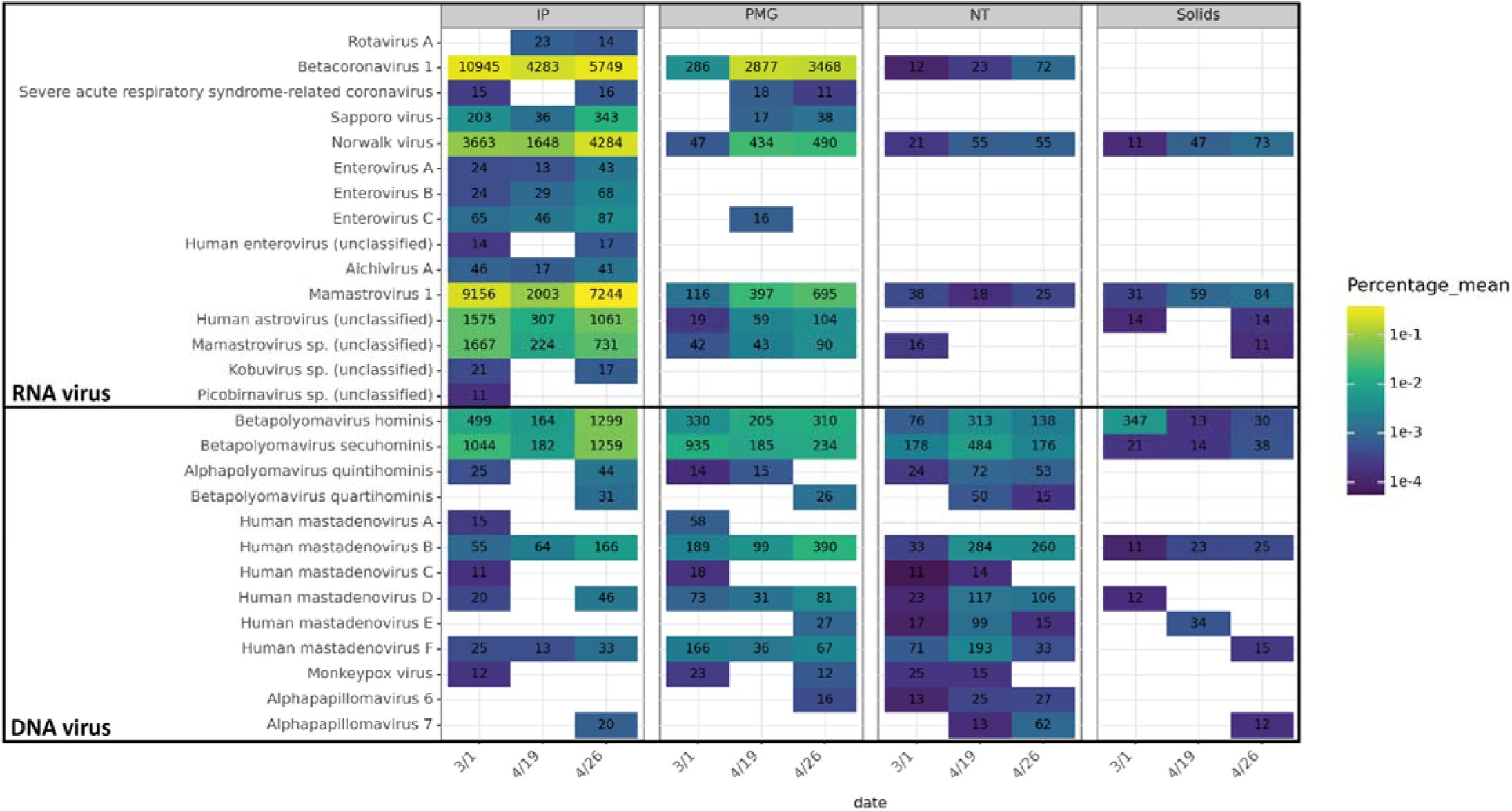
Relative abundance of human virus species in each sample. Fill indicates the average percent relative abundance of each virus species in total unique reads across triplicate samples, based on Recentrifuge read classification. Species with fewer than an average of 10 reads per sample are not shown. Text in each cell indicates the average read counts assigned to the species for each sample. Viruses are grouped by genome type. NCBI taxIDs corresponding to species without names (e.g. “sp.”) are appended with “(unclassified)” (see supplementary methods). Note that Betacoronavirus 1 includes the spike-in bovine coronavirus.

### 3.4 Potential of recovering near-complete human virus genomes

Seven near-complete human virus genomes were assembled from IP samples, the most from any concentration/extraction method (Figure S4b). This aligned with the high numbers of total virus and human virus reads in these samples (59,965 ± 28,180 and 20,242 ± 9,294, respectively, Table S1). No near-complete human virus genomes were obtained from Solids-extracted samples (Figure S4b) likely due to insufficient reads for total viruses and human viruses (11,043 ± 2,720 and 213 ± 99, respectively, Table S1). These results highlight the need to understand the minimum sequencing depth in relation to the proportion of viral reads required for the assembly of high-quality virus genomes.

JC polyomavirus composite genomes were assembled in samples from three concentration/extraction methods (IP, PMG, and NT) and multiple replicates (Figure S4b). The recovery of JC polyomavirus genomes is perhaps unsurprising given that approximately 40% of the population sheds the virus through urine ^54^. Also, as a non-enveloped DNA virus with a circular genome, JC polyomavirus is highly resistant to environmental stress and exonuclease activity ^9^. Ten scaffolds classified as near-complete JC polyomavirus genomes were used for phylogenetic analysis. At least one subtype of JC polyomavirus 3 was present (Node 1353 NT_301_1), affiliated with clades from South Africa (Figure S7). Although other scaffolds were clustered together, they exhibited relatively low node support values (< 50); likely several of these scaffolds represent the same JC polyomavirus population in replicate wastewater samples, with variations in the composite assembly. These results, and those from other recent studies ^25^, demonstrated that probe capture enrichment can yield whole genomes of high-abundance viruses for phylogenetic analysis, which may be useful for identifying novel virus strains in the future.

### 3.5 The comparison of SARS-CoV-2 detection between dPCR and targeted sequencing using the VSP panel

To compare the sensitivity of sequencing to that of digital PCR, endogenous SARS-CoV-2 and the spike-in BCoV were quantified in the final extracted nucleic acids produced by each concentration method. By sequencing, both SARS-CoV-2 and BCoV were detected in PMG and IP samples at the employed alignment stringency and read count threshold (> 10 reads, Figure 5), which corresponded with the higher relative abundances of human viruses in these two methods. However, the absolute concentrations of SARS-CoV-2 were significantly lower in IP samples than in PMG samples (IP v.s. PMG p = 0.009, Figure 5a and Table S6). Target virus concentrations could be increased by increasing the effective volume of wastewater processed. Specifically, the final volume of the ultrafilter concentrate nearly always exceeded the maximum input for nucleic acid extraction, resulting in a lower effective volume (ranging from 16.3 ± 13 mL, Table S1). Similarly, only 0.25 g was extracted from 0.60 ± 0.18 g of wet solids due to the limitation of the extraction kit, resulting in a lower effective sample volume processed relative to the PMG and NT methods. The limited input may partially explain the low concentrations of targets observed by dPCR. Notably, although samples from March 1 showed similar SARS-CoV-2 concentrations from both NT and PMG methods (NT: 14.5 ± 3.4 gc/µL, PMG: 9.2 ± 1.4 gc/µL, p = 0.18, Table S6), no SARS-CoV-2 reads were detected in the NT samples from this date. Meanwhile, although BCoV was detected by dPCR in NT and Solids samples at low levels, it was absent in the sequencing results. This suggests that in addition to the absolute viral concentration indicated by dPCR, background non-target sequences may also influence target detection by sequencing.

**Figure 5.**
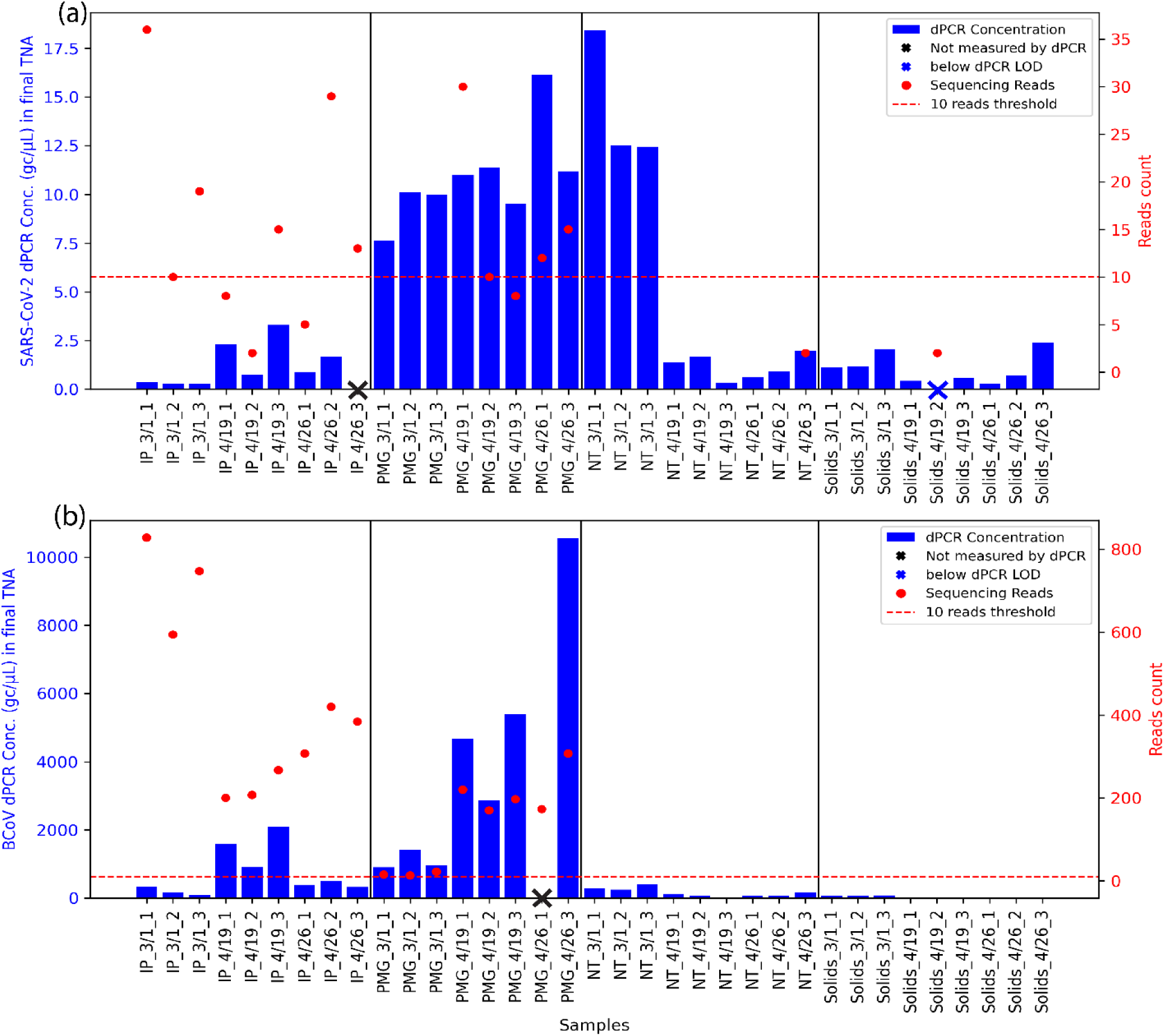
Detection sensitivity comparison between dPCR and reads-based classification (Recentrifuge) of sequencing results. (a) SARS-CoV-2 detection comparison; (b) BCoV detection comparison. Blue bars on the left y-axis represent the virus concentration measured by dPCR in the final total nucleic acids (TNA) eluted in 100 µl after each extraction. Samples with dPCR concentration below the operational limit of detection are shown with a blue “x”, and samples without measurement were labeled with a black “x”. Red points on the right y-axis represent virus read counts from unique reads. The dashed red line at 10 reads indicates the operational limit of detection used elsewhere in the analysis.

### 3.6 Implications for genome surveillance of known and novel human viruses

Based on the comparisons reported here, wastewater virus concentration/extraction methods should be chosen carefully and aligned with the specific monitoring endpoint and goal (e.g., sequencing or dPCR, specific targets or broad range of targets). Removing wastewater solids, after treatment with either Tween 20 (IP method) or protease (PMG method) and prior to concentration and extraction, improved the overall detection of human viruses in probe-capture sequencing by minimizing the ratio between off-target sequences and targeted human virus sequences ^58^ (Figure 2). However, solids removal may also decrease the sensitivity of virus detection by dPCR by decreasing the absolute quantities of the target in the sample (Figure 5) ^18^. As untargeted sequencing was not performed in parallel, the extent to which solids removal improved probe-capture enrichment specifically cannot be directly quantified. Given that methods that included solids showed higher relative abundances of DNA viruses, a DNase treatment may improve the recovery of human RNA viruses with these methods. Additionally, while only two extraction methods were applied here (Qiagen AllPrep PowerViral and Promega Wizard Enviro TNA), the extraction method used for solids and viral concentrates may affect the overall sensitivity of sequencing by influencing the degree of viral lysis and integrity of the resulting nucleic acids ^16^. Further studies should be performed during periods of higher target concentration in wastewater (e.g., SARS-CoV-2 surges) or using spike-in viruses to quantitatively determine limits of detection for sequencing using different concentration and extraction methods.

Finally, the choice of probe panel likely also impacts the sensitivity of virus detection in probe-capture sequencing. When using the RVOP probe set (which contains fewer virus targets than the VSP probe set) several studies found remarkably high coverages of SARS-CoV-2, surpassing that of other human viruses included in the RVOP panel ^14, 15, 49^. However, in the present study and other studies using broad virus capture panels ^23, 48^, sequence data were dominated by enteric viruses such as mamastrovirus, with limited detection of SARS-CoV-2. This points to the inherent challenge of using broad panels as a means of wastewater-based surveillance for early detection of novel virus strains, which may appear at low abundance before going on to cause a larger outbreak.

## Supporting information

Supplementary tables included in the supplementary materials

## Data Availability

All data produced in the present study are available upon reasonable request to the authors

https://github.com/mj2770/Wastewater-virus-surveillance-

## Acknowledgements

Funding was provided by the UCOP Lab Fees CRT Award (L22CR4507). We thank Khi Lai at EBMUD for sample collection and Sanaiya Islam for laboratory management. Library preparation was performed with advice from Justin Choi and Byran Bach at the Functional Genomics Laboratory and sequencing was performed at the Vincent J. Coates Sequencing Laboratory (QB3, UC Berkeley, RRID: SCR_022170). We thank Allie Nguyen and Van Trinh for their assistance with laboratory and bioinformatic analyses.

