## Supplementary materials for "Evaluation of the impact of concentration and extraction methods on the targeted sequencing of human viruses from wastewater"

#### Supplementary Tables:

**Table S1.** All 36 samples' processing includes concentration/extraction, quality control, and raw sequencing data QC trimming/deduplication statistics

**Table S2.** (a) Primers, probes, and cycling parameters for RT-dPCR quantification; (b) Reaction mixtures for 8.5k and 26k 24-well nanoplates

**Table S3.** dMIQE checklist for RT-dPCR experiments

**Table S4.** GISAID SARS-CoV-2 reference genome accession numbers

**Table S5.** 66 virus targets of high public health significance in the Illumina VSP panel

**Table S6.** Comparison of dPCR and sequencing reads-based classification of SARS-CoV-2 and BCoV

#### Supplementary Figures:

**Figure S1.** Partition fluorescence plots of positive and negative control

**Figure S2.** Clustering of all samples by PCoA plot based on the calculated MASH distance of virus sequences classified by Centrifuge

**Figure S3.** (a) The richness of detected virus species at the species level; (b) The relative abundance of the detected viruses included in the VSP panel

**Figure S4**. (a) Assessment of assembly quality based on N50 and total assembly length; (b) Count of near-complete virus genomes assembled

**Figure S5.** Representative assembly visual inspection by Integrative Genomics Viewer (IGV)

**Figure S6.** Dotplots of assembled putative JC polyomavirus scaffolds with repeated regions at the beginning and the end of the sequence

**Figure S7.** Maximum likelihood phylogenetic tree of assembled JC polyomavirus scaffolds

#### Supplementary Methods

- Validation of virus-host classification
- DNA and RNA virus classification
- Human virus species classification

**Table S1:** See the Excel file

**Table S2:**

(a) Primers, probes, and cycling parameters for SARS-CoV-2 dPCR quantification

| Targeted Virus | Assays | Oligonucleotides sequences (5' - 3') (purchased from IDT) | Cycling parameters | References |
|---|---|---|---|---|
| SARS-CoV-2 | N1 | F: GACCCCAAAATCAGCGAAAT<br>R: TCTGGTTACTGCCAGTTGAATCTG<br>P: FAM-ACCCCGCATTACGTTTGGTGGACC | 50 °C for 40 min for RT; 95 °C for 2 min; 45 cycles of 95 °C for 5 s; 60 °C for 30 s | US CDC, 2020 |
| Bovine coronavirus | BCoV | F: CTGGAAGTTGGTGGAGTT<br>R: ATTATCGGCCTAACATACATC<br>P: HEX-CCTTCATATCTATACACATCAAGTTGTT | | Decaro, 2008* |

* Decaro, Nicola et al. "Detection of bovine coronavirus using a TaqMan-based real-time RT-PCR assay." Journal of virological methods vol. 151,2 (2008): 167-171. doi:10.1016/j.jviromet.2008.05.016

(b) Reaction mixtures for 8.5k and 26k 24 wells nanoplates

| Plate types | Total reaction volumes (μL) | Mastermix (μL) | Primer and probe mix (μL) 400 nM of forward primer, 400 nM of reverse primer, 200 nM of probe | Nuclease free water (μL) | Samples/negative controls/standards (μL) | Reverse transcriptase (μL) |
|---|---|---|---|---|---|---|
| 8.5k 24 wells | 12 | 3 | 0.6 | 5.28 | 3 | 0.12 |
| 26k 24 wells | 40 | 10 | 2 | 17.6 | 10 | 0.4 |

**Table S3:** dMIQE checklist for RT-dPCR experiments

| ITEM TO CHECK | PROVIDED | COMMENT |
|---|---|---|
| **1. SPECIMEN** | | |
| Detailed description of specimen type and numbers | **Y** | See Methods |
| Sampling procedure (including time to storage) | **Y** | See Methods |
| Sample aliquotation, storage conditions and duration | **Y** | See Methods |

| | | |
|---|---|---|
| **2. NUCLEIC ACID EXTRACTION** | | |
| Description of extraction method including amount of sample processed | **Y** | See Methods |
| Volume of solvent used to elute/resuspend extract | **Y** | See Methods |
| Number of extraction replicates | **Y** | See Methods |
| Extraction blanks included? | **Y** | See Methods |
| **3. NUCLEIC ACID ASSESSMENT AND STORAGE** | | |
| Method to evaluate quality of nucleic acids | **Y** | See Methods |
| Method to evaluate quantity of nucleic acids (including molecular weight and calculations when using mass) | **Y** | See Methods |
| Storage conditions: temperature, concentration, duration, buffer, aliquots | **Y** | See Methods |
| Clear description of dilution steps used to prepare working DNA solution | **Y** | See Methods |
| **4. NUCLEIC ACID MODIFICATION** | | |
| Template modification (digestion, sonication, pre-amplification, bisulphite etc.) | **N** | N/A |
| Details of repurification following modification if performed | **N** | N/A |
| **5. REVERSE TRANSCRIPTION** | | |
| cDNA priming method and concentration | **Y** | See Methods |
| One or two step protocol (include reaction details for two step) | **Y** | See Methods |
| Amount of RNA added per reaction | **Y** | See Methods |
| Detailed reaction components and conditions | **Y** | See Methods |
| Estimated copies measured with and without addition of RT* | **N** | No need |
| Manufacturer of reagents used with catalogue and lot numbers | **Y** | See Methods |
| Storage of cDNA: temperature, concentration, duration, buffer and aliquots | **N** | N/A |
| **6. dPCR OLIGONUCLEOTIDES DESIGN AND TARGET INFORMATION** | | |
| Sequence accession number or official gene symbol | **Y** | Table S2 |
| Method (software) used for design and in silico verification | **N** | N/A |
| Location of amplicon | **N** | N/A |

| | | |
|---|---|---|
| Amplicon length | **Y** | Table S2 |
| Primer and probe sequences (or amplicon context sequence)** | **Y** | Table S2 |
| Location and identity of any modifications | **N** | N/A |
| Manufacturer of oligonucleotides | **Y** | Table S2 |
| **7. dPCR PROTOCOL** | | |
| Manufacturer of dPCR instrument and instrument model | **Y** | See Methods |
| Buffer/kit manufacturer with catalogue and lot number | **Y** | See Methods |
| Primer and probe concentration | **Y** | See Methods |
| Pre-reaction volume and composition (incl. amount of template and if restriction enzyme added) | **Y** | See Methods |
| Template treatment (initial heating or chemical denaturation) | **N** | N/A |
| Polymerase identity and concentration, Mg++ and dNTP concentrations*** | **N** | N/A |
| Complete thermocycling parameters | **Y** | Table S2 |
| **8. ASSAY VALIDATION** | | |
| Details of optimisation performed | **N** | N/A |
| Analytical specificity (vs. related sequences) and limit of blank (LOB) | **N** | N/A |
| Analytical sensitivity/LoD and how this was evaluated | **Y** | See Methods |
| Testing for inhibitors (from biological matrix/extraction) | **N** | N/A |
| **9. DATA ANALYSIS** | | |
| Description of dPCR experimental design | **Y** | See Methods |
| Comprehensive details negative and positive of controls (whether applied for QC or for estimation of error) | **Y** | See Methods |
| Partition classification method (thresholding) | **Y** | See Methods |
| Examples of positive and negative experimental results (including fluorescence plots in supplemental material) | **Y** | Figure S1 |
| Description of technical replication | **N** | No technical replicates |
| Repeatability (intra-experiment variation) | **N** | N/A |
| Reproducibility (inter-experiment/user/lab etc. variation ) | **N** | N/A |

| | | |
|---|---|---|
| Number of partitions measured (average and standard deviation ) | **Y** | See Methods |
| Partition volume | **N** | N/A |
| Copies per partition (λ or equivalent ) (average and standard deviation) | **N** | N/A |
| dPCR analysis program (source, version) | **Y** | See Methods |
| Description of normalisation method | **N** | N/A |
| Statistical methods used for analysis | **Y** | See Methods |
| Data transparency | raw data available on request: | |

**Table S4:** See the Excel file

**Table S5:** 66 targeted viruses included in the Illumina VSP panel. The Illumina website does not provide a complete list of 203 strains under these 66 viruses, and the naming convention is a mix of genus, species, and sub-species levels. To track the detection of targeted viruses by each concentration/extraction method, the 66 virus names were manually checked against the NCBI taxonomy. Exact matches were recorded with virus name and taxID. For names not exactly matching NCBI taxonomy, potentially included viruses were recorded with NCBI names, taxID, and ranks under the Illumina virus names.

| Illumina Name | NCBI Name | taxID | Rank | Symptom type |
|---|---|---|---|---|
| Adenovirus | Mastadenovirus | 10509 | genus | Respiratory/Cardiopulmonary |
| Aichivirus | Aichivirus A | 72149 | species | Enteric |
| | Aichivirus B | 194965 | species | Enteric |
| | Aichivirus C | 1298633 | species | Enteric |
| | Aichivirus D | 1897731 | species | Enteric |
| | Aichivirus E | 1986958 | species | Enteric |
| | Aichivirus F | 1986959 | species | Enteric |
| Astrovirus | Astroviridae | 39733 | family | Enteric |
| Chapare virus | Chapare virus | 3052302 | species | Hemorrhagic fever |
| Chikungunya virus | Chikungunya virus | 37124 | species | Anthropod tranmitted/febrile tropical |
| Coronavirus-229E | Coronavirus 229E | 11137 | species | Respiratory/Cardiopulmonary |
| Coronavirus-HKU1 | Human coronavirus HKU1 | 290028 | species | Respiratory/Cardiopulmonary |

| | | | | |
|---|---|---|---|---|
| Coronavirus-OC43 | Human coronavirus OC43 | 31631 | no rank | Respiratory/Cardiopulmonary |
| Coronavirus-NL63 | Coronavirus NL63 | 277944 | species | Respiratory/Cardiopulmonary |
| Coxsackievirus | Coxsackievirus | 12066 | species | Enteric |
| | Coxsackievirus A9 | 12067 | no rank | Enteric |
| | Coxsackievirus A22 | 42783 | no rank | Enteric |
| | Coxsackievirus A19 | 42778 | no rank | Enteric |
| | Coxsackievirus A1 | 42779 | no rank | Enteric |
| | Coxsackievirus A4 | 42785 | no rank | Enteric |
| | Coxsackievirus B4 | 12073 | no rank | Enteric |
| | Coxsackievirus B5 | 12074 | no rank | Enteric |
| | Coxsackievirus A6 | 86107 | no rank | Enteric |
| Crimean-congo haemorrhagic fever virus | Crimean-congo haemorrhagic fever virus | 3052518 | species | Hemorrhagic fever |
| Dengue virus 1 | Dengue virus 1 | 11053 | no rank | Anthropod tranmitted/febrile tropical |
| Dengue virus 2 | Dengue virus 2 | 11060 | no rank | Anthropod tranmitted/febrile tropical |
| Dengue virus 3 | Dengue virus 3 | 11069 | no rank | Anthropod tranmitted/febrile tropical |
| Dengue virus 4 | Dengue virus 4 | 11070 | no rank | Anthropod tranmitted/febrile tropical |
| Eastern equine encephalitis virus | Eastern equine encephalitis virus | 11021 | species | Encephalitis |
| Ebola virus | Ebola virus | 1570291 | no rank | hemorrhagic fever |
| Enterovirus | Enterovirus | 12059 | genus | Enteric |
| Guanarito virus | Guanarito virus | 3052307 | species | hemorrhagic fever |
| Hantavirus | Hantavirus | 1980442 | genus | Respiratory/Cardiopulmonary |
| Hendra henipavirus | Hendra henipavirus | 3052223 | species | Encephalitis |
| Hepatitis A virus | Hepatitis A virus | 12092 | species | Bloodborne |
| Hepatitus B virus | Hepatitis B virus | 10407 | species | Bloodborne |
| Hepatitus C virus | Hepatitis C virus | 3052230 | species | Bloodborne |
| Hepatitus E virus | Hepatitis E virus/Hepeviridae | 291484 | family | Bloodborne |
| Human Immunodeficiency Virus 1 | Human Immunodeficiency Virus 1 | 11676 | species | Bloodborne |

| | | | | |
|---|---|---|---|---|
| Human Immunodeficiency Virus 2 | Human Immunodeficiency Virus 2 | 11709 | species | Bloodborne |
| Influenza A Virus | Influenza A Virus | 11320 | no rank | Respiratory/Cardiopulmonary |
| Influenza B Virus | Influenza B Virus | 11520 | no rank | Respiratory/Cardiopulmonary |
| Japanese encephalitis virus | Japanese encephalitis virus | 11072 | no rank | Encephalitis |
| Junin virus | Junin virus | 2169991 | species | hemorrhagic fever |
| Kyasanur Forest disease virus | Kyasanur Forest disease virus | 33743 | no rank | hemorrhagic fever |
| Lassa fever virus | Lassa mammarenavirus | 3052310 | species | hemorrhagic fever |
| Lujo hemorrphagic fever virus | Lujo mammarenavirus | 3052314 | species | hemorrhagic fever |
| Machupo virus | Machupo mammarenavirus | 3052317 | species | hemorrhagic fever |
| Marburg virus | Marburg marburgvirus | 3052505 | species | hemorrhagic fever |
| MERS-CoV | MERS coronavirus | 1335626 | species | Respiratory/Cardiopulmonary |
| Metapneumovirus | Metapneumovirus | 162387 | genus | Respiratory/Cardiopulmonary |
| Monkeypox virus | Monkeypox virus | 10244 | species | Rash/lesion |
| Nipah virus | Nipah virus | 3052225 | species | Encephalitis |
| Norovirus | Norovirus | 142786 | genus | Enteric |
| Omsk hemorrhagic fever virus | Omsk hemorrhagic fever virus | 12542 | no rank | hemorrhagic fever |
| Parainfluenza virus | Human parainfluenza virus Hue-2015 | 2559896 | species | Respiratory/Cardiopulmonary |
| | Human parainfluenza virus HCM-2015 | 2559897 | species | Respiratory/Cardiopulmonary |
| | Human respirovirus 1 | 12730 | no rank | Respiratory/Cardiopulmonary |
| | Human orthorubulavirus 2 | 2560525 | no rank | Respiratory/Cardiopulmonary |
| | Human respirovirus 3 | 11216 | no rank | Respiratory/Cardiopulmonary |
| | Human parainfluenza virus 4a | 11224 | no rank | Respiratory/Cardiopulmonary |
| | Human parainfluenza virus 4b | 11226 | no rank | Respiratory/Cardiopulmonary |
| | Parainfluenza virus 5 | 2905673 | no rank | Respiratory/Cardiopulmonary |

| | | | | |
|---|---|---|---|---|
| Parechovirus | Parechovirus | 138954 | genus | Respiratory/Cardiopulmonary |
| Oncolytic human papillomavirus | human papillomavirus | 10566 | species | Oncogenic |
| Parvovirus | Parvovirus | 1506574 | genus | Respiratory/Cardiopulmonary |
| Poliovirus | Human poliovirus 1 | 12080 | serotype | Enteric |
| | Human Poliovirus type 2 | 12083 | serotype | Enteric |
| | Human poliovirus 3 | 12086 | serotype | Enteric |
| | wild poliovirus type 3 | 53259 | serotype | Enteric |
| | recombinant polioviruses | 909390 | no rank | Enteric |
| | Polyomavirus sp. | 36362 | species | Oncogenic |
| Respiratory syncytial virus | Respiratory syncytial virus | 12814 | no rank | Respiratory/Cardiopulmonary |
| Rhinovirus | Rhinovirus A | 147711 | species | Respiratory/Cardiopulmonary |
| | Rhinovirus B | 147712 | species | Respiratory/Cardiopulmonary |
| | Rhinovirus C | 463676 | species | Respiratory/Cardiopulmonary |
| | unclassified rhinovirus | 348531 | no rank | Respiratory/Cardiopulmonary |
| Rift Valley fever virus | Rift Valley fever virus | 11588 | no rank | hemorrhagic fever |
| Rotavirus | Rotavirus | 10912 | genus | Enteric |
| Rubella virus | Rubella virus | 11041 | no rank | Rash/lesion |
| Sabia virus | Sabia virus | 2907957 | no rank | hemorrhagic fever |
| Salivirus | Salivirus | 688449 | genus | Enteric |
| Sapovirus | Sapovirus | 95341 | genus | Enteric |
| SARS-COV | Severe acute respiratory syndrome coronavirus | 2901879 | no rank | Respiratory/Cardiopulmonary |
| SARS-COV-2 | Severe acute respiratory syndrome coronavirus 2 | 2697049 | no rank | Respiratory/Cardiopulmonary |
| Tick-borne encephalitis virus | Tick-borne encephalitis virus | 11084 | no rank | Encephalitis |
| Torque Teno virus | Torque Teno virus | 68887 | species | Bloodborne |
| Variola virus | Variola virus | 10255 | species | Rash/lesion |
| Venezuelan equine encephalitis virus | Venezuelan equine encephalitis virus | 11036 | species | Encephalitis |
| West Nile virus | West Nile virus | 11082 | no rank | Anthropod tranmitted/febrile tropical |

| | | | | |
|---|---|---|---|---|
| Western equine encephalitis virus | Western equine encephalitis virus | 11039 | species | Encephalitis |
| Yellow fever virus | Yellow fever virus | 11089 | no rank | Anthropod tranmitted/febrile tropical |
| Zika virus | Zika virus | 64320 | no rank | Anthropod tranmitted/febrile tropical |

**Table S6.** See the Excel file

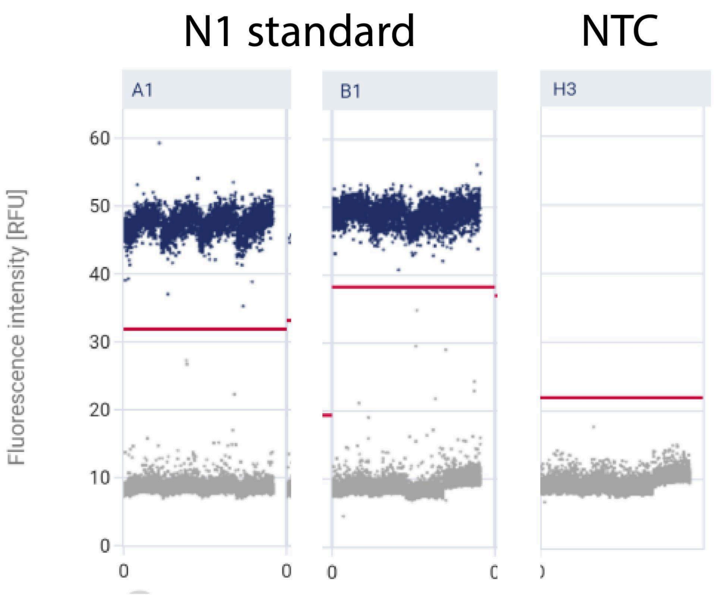


**Figure S1.** Examples of partition fluorescence plots of dPCR positive and negative control for the CDC N1 assay for SARS-CoV-2.

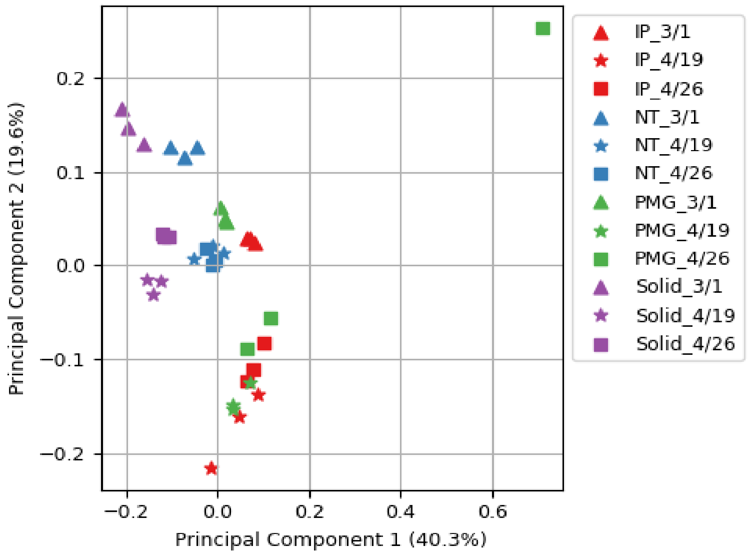


**Figure S2.** Clustering of all samples by PCoA plot based on the calculated MASH distance of virus sequences classified by Centrifuge. PMG_4/26_2 sample exhibited distinct properties as compared to the other two biological replicates.

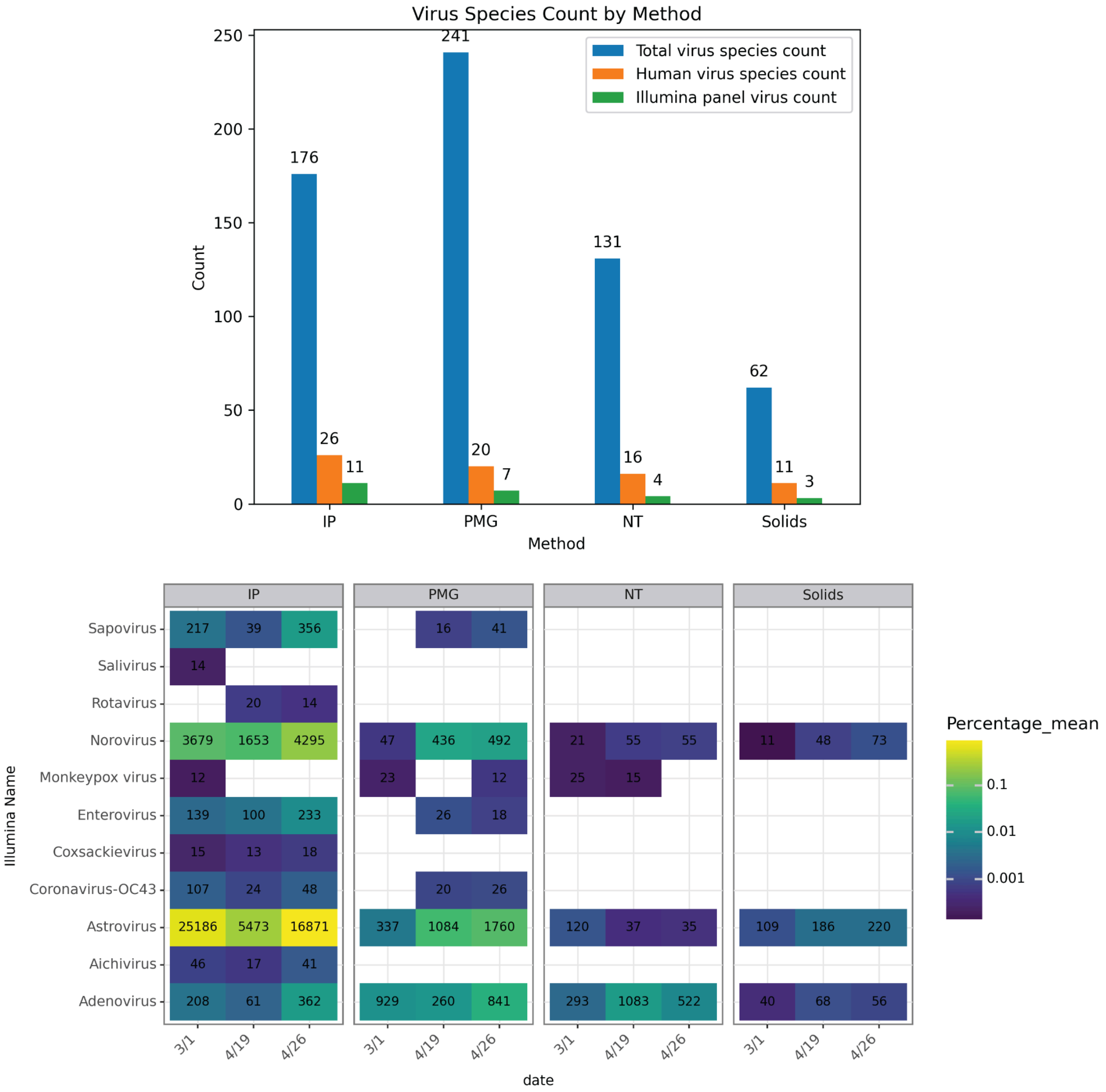


**Figure S3.** (a) The richness of detected virus species at the species level, encompassing total viruses, human viruses, and those targeted by the Illumina VSP panel. The richness was calculated by counting the total unique taxIDs assigned to species levels in each method; (b) The percentages of the detected viruses included in the VSP panel in total unique reads, filtered by counts exceeding 10 reads. Text in each cell indicates the average read counts assigned to the virus for each sample.

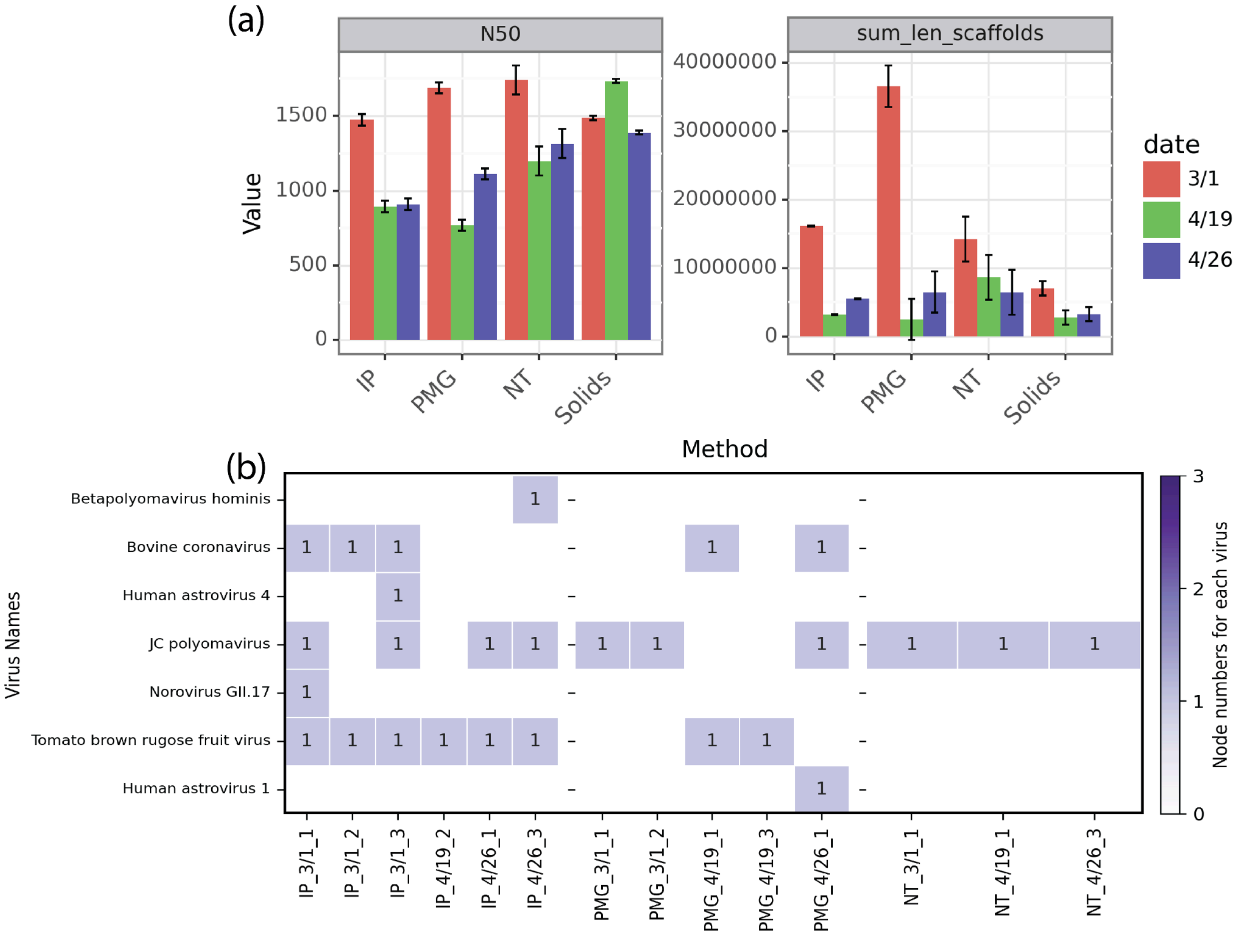


**Figure S4**. (a) Assessment of assemblies from each method. The assembly quality is based on N50 and the total assembly length. (b) Count of near-complete virus genomes assembled after applying strict criteria (>1000 bp, >10 average coverage depth, >70% coverage breadth of the complete genome, >80% identity, >90% alignment/query length, best hits for each scaffold, and matching complete genomes) to both scaffolds and BLASTn results. Samples with no recovered near-complete virus genomes are not shown.

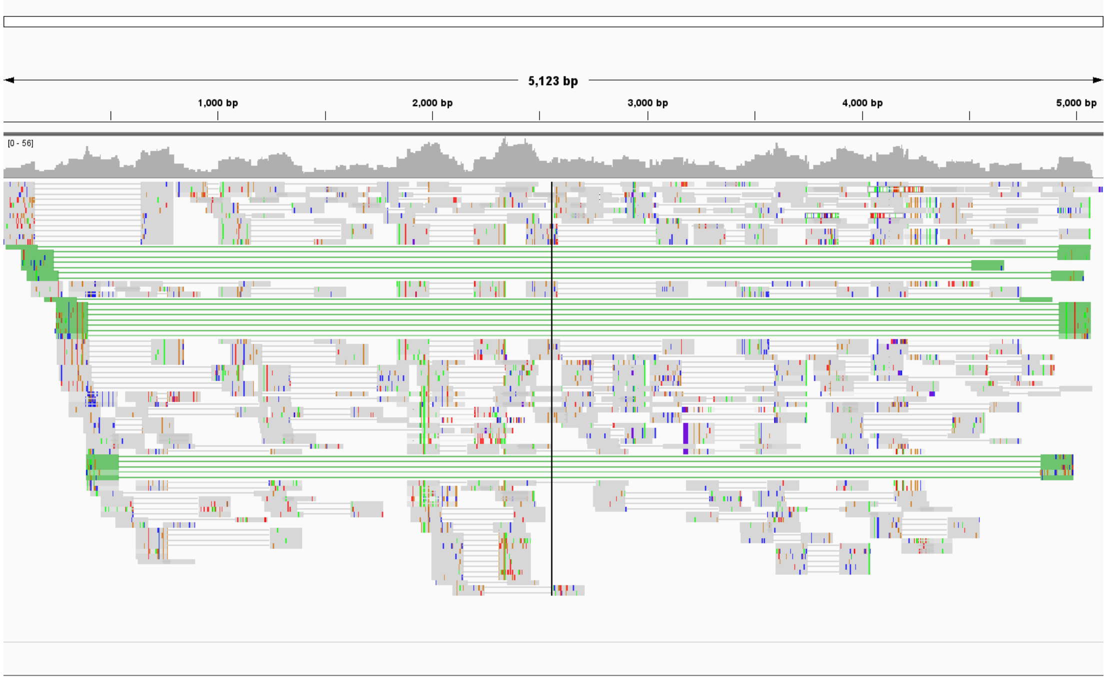


**Figure S5.** Representative assembly visual inspection by Integrative Genomics Viewer (IGV). The reads were mapped to the assembled putative JC polyomavirus scaffolds (NODE_58_length_5177_cov_12.765912||full) from the PMG_426_1 sample. Both coverage and alignment tracks were shown with mismatches. All reads were colored by pair orientation and shaded by mapping quality high. The coverage allele frequency threshold was set to 0.2. Note that the genome is circular, resulting in read pairs with mates mapping to the 5' and 3' ends when viewed linearly.

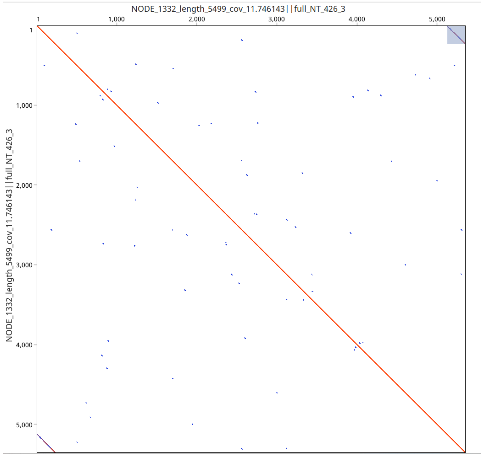


**Figure S6.** Dotplots of assembled putative JC polyomavirus scaffolds with repeated regions at the beginning and the end of the sequence (see gray box selected in upper right corner). Plot shown represents the sequence of NODE_1332_length_5499_cov_11.746143||full from NT_426_3 sample.

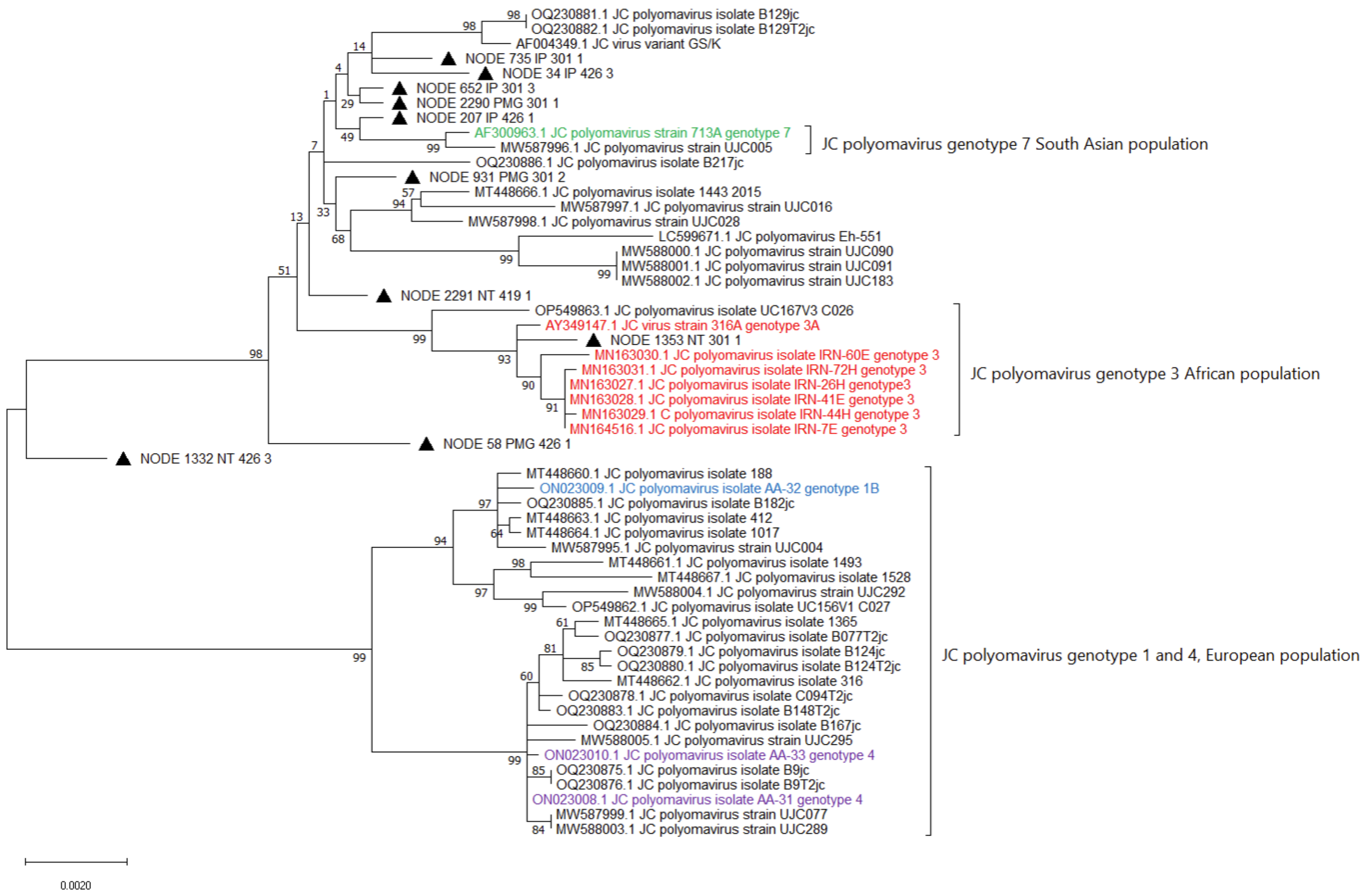


**Figure S7**. **Maximum likelihood phylogenetic tree of assembled JC polyomavirus genomes**. The tree was generated using the Maximum Likelihood method and the Tamura-Nei model. Node support values, indicating the percentage of trees in which associated taxa clustered together, were obtained from 100 bootstraps. This analysis included 56 unique nucleotide sequences, with 4168 columns in the final alignment. Reference genomes were color-coded based on the NCBI database, with genomes labeled according to their subtypes or types (see detailed information below). Scaffolds from wastewater assemblies are indicated with triangles.

AF300963.1: Cui,X. and Stoner,G.L. Direct Submission, submitted (29-AUG-2000) NINDS, Neurotoxicology Section, National Institutes of Health, 36 Convent Drive, Bldg. 36, Room 4A27, Bethesda, MD 20892, USA, https://www.ncbi.nlm.nih.gov/nuccore/AF300963.1

AY349147.1: Mengistu,G. and Stoner,G.L. Direct Submission, submitted (23-JUL-2003) National Institute of Neurological Disorders and Stroke, National Institutes of Health, 36 Convit Dr., Bldg. 36, Rm3A-11, Bethesda, MD 20892, USA, https://www.ncbi.nlm.nih.gov/nuccore/AY349147.1

MN163027.1-MN163031.1: Pirmorad, R. and Makvandi,M. Direct Submission, submitted (10-JUL-2019) medical science department, Ahvaz Jundishapour University of Medical Science, Golestan district-Esfand Street, Ahvaz, Khuzestan 61335, Iran, https://www.ncbi.nlm.nih.gov/nuccore/MN163030.1

MN164516.1: Pirmorad, R. and Makvandi, M. Direct Submission, submitted (04-JUL-2019) medical science department, Ahvaz Jundishapour University of Medical Science, Golestan district-Esfand Street, Ahvaz, Khuzestan 61335, Iran, https://www.ncbi.nlm.nih.gov/nuccore/MN164516.1

ON023008.1-ON023010.1: Lari Pyöriä, Diogo Pratas, Mari Toppinen, Klaus Hedman, Antti Sajantila, Maria F Perdomo, Unmasking the tissue-resident eukaryotic DNA virome in humans, Nucleic Acids Research, Volume 51, Issue 7, 24 April 2023, Pages 3223–3239, https://doi.org/10.1093/nar/gkad199

### Supplementary methods

**Validation of virus-host classification**: Upon manual inspection of the resulting host assignments, reads classified as bacteriophage sp. (taxID 38018) and uncultured phage (taxID 278008) were found to have host assigned as “bacteria|human|invertebrates|vertebrates” and “bacteria|fungi|human|invertebrates”, respectively. The host for these reads was re-assigned as “bacteria”. Additionally, the uncultured viruses (taxID 340016) and circular genetic element sp. (taxID 2202954) were originally assigned to “bacteria|human|invertebrates|land plants|protozoa” and “invertebrates|vertebrates”, respectively. The host for these reads was re-assigned as “unknown”. Lastly, reads classified as bovine coronavirus were assigned within NCBI as corresponding to the “human|vertebrate” host category and were removed before virus-host analysis.

**DNA and RNA virus classification**: Six virus kingdoms, namely Orthornavirae, Heunggongvirae, Loebvirae, Sangervirae, Shotokuvirae, and Bamfordvirae, were identified across all methods. These kingdoms were further classified into RNA (Orthornavirae), double-strand DNA (Bamfordvirae and Heunggongvirae), and single-strand DNA viruses (Loebvirae, Sangervirae, Shotokuvirae), following the realms outlined by the International Committee on Taxonomy of Viruses (ICTV).

**Human virus species classification**: The unclassified species includes Human enterovirus (taxID: 1193974), Kobuvirus sp. (taxID: 2094719), Picobirnavirus sp. (taxID: 1907787), Mamastrovirus sp. (taxID: 1912147), Human astrovirus. (taxID:1868658). To denote that these may not represent true species but rather a combination of several unclassified species assigned to the same taxID in NCBI, “unclassified” was appended to the species name.
